# Displaced by a Volcano: Adolescent Well-Being in Uncertain Times in Iceland, 2023–2026

**DOI:** 10.64898/2026.11.09.26367581

**Authors:** David Reimer, Juuso Repo, Elina Kilpi-Jakonen, Kolbrún Þ. Pálsdóttir

## Abstract

**Aims:** The November 2023 evacuation of Grindavík, Iceland, left families facing prolonged uncertainty about return. We examined adolescent well-being before and after evacuation and changes in gaps with national peers between 2025 and 2026.

**Methods:** Repeated cross-sectional analyses used the 2023, 2025, and 2026 Icelandic Youth Study waves, adjusting for grade, gender, and non-Icelandic background. The town’s single school in 2023 provided the pre-evacuation benchmark, with models accounting for national time trends. Adolescents who moved before evacuation provided a mobility benchmark.

**Results:** In 2025, evacuees reported lower life satisfaction and school belonging, more psychosomatic symptoms, and higher odds of school absence and past-30-day substance use than the national reference group. The three well-being outcomes departed from the pre-evacuation pattern. Life-satisfaction and belonging gaps were larger among girls than boys in 2025, but these gender differences narrowed by 2026. Comparisons with prior movers suggested a distinct evacuation-related pattern in 2025 among evacuee girls, who reported lower life satisfaction and school belonging than prior-mover girls. In 2026, psychosomatic symptoms (B = 0.29, 95% CI [0.08, 0.49]), absenteeism (OR = 1.81, 95% CI [1.26, 2.60]), and substance use (OR = 2.59, 95% CI [1.75, 3.84]) remained higher relative to the national reference group. Life satisfaction and belonging were directionally lower, with intervals including zero. Change estimates did not establish narrowing of the gaps.

**Conclusions:** The findings underscore the need for sustained educational and well-being support to address adolescents’ social connections alongside their mental health during prolonged displacement, including in a Nordic welfare setting.

## Introduction

Natural disasters that unfold slowly can leave communities in prolonged uncertainty about whether, when, and how residents can or should return home. The town of Grindavík on Iceland’s Reykjanes Peninsula is such a case. After several years of seismic unrest and nearby eruptions, Grindavík was evacuated on November 10^th^ 2023 as magma moved beneath the town causing major earthquakes (Parks et al., 2024). All 3,790 residents had to leave, and repeated eruptions over the following months made a return unlikely for most families (Alþingi, 2024). For more than two years, the civil protection authorities rated the town as high risk for families with children, before lowering the risk level in mid-year 2026 (Almannavarnir, 2026). In the immediate aftermath of the town’s evacuation, authorities prioritised the immediate well-being and security of Grindavík residents. Considerable emphasis was also on providing social and psychological support, not least for children and young people (Alþingi, 2024). Schooling continued, at first partly in temporary Grindavík classes organized in the capital area and, from June 2024, in local schools across the country. By fall 2024, Grindavík children were spread across 73 schools around Iceland, though the majority of Grindavík families had relocated either to the capital area or nearby municipalities on the Reykjanes peninsula (Framkvæmdanefnd Grindavíkur, 2025; for details, see Reimer et al., 2026). The town’s school reopened in August 2026 with about 30 students, with most families remaining elsewhere (Víkurfréttir, 2026). For adolescents, the evacuation meant losing a school community and neighbourhood friendships, moving home repeatedly, and settling into new schools, without knowing whether the move was temporary or permanent. In the first five months after the evacuation, families changed homes about three times on average (Hrólfsson, 2024).

These disruptions occurred in a Nordic welfare setting which supported displaced families financially and provided access to education and social services. The government-appointed Grindavík Committee coordinated support for evacuated families and for the schools that received Grindavík students; its report described concern for emotional and social difficulties among students in host schools (Framkvæmdanefnd Grindavíkur, 2025). Among adults, a 2025 government survey found that 66% of evacuees reported worse mental health than before the evacuation and that 46% still regarded their housing as temporary (Forsætisráðuneytið, 2025).

Systematic evidence on the Grindavík adolescents comes so far from the 2025 wave of the Icelandic Youth Study, in which evacuees reported lower well-being than their peers nationally (Reimer et al., 2026). Like the reports above, it captures a single point in time and cannot show whether such differences are a transient reaction or the start of lasting difficulties. The present study adds a pre-evacuation benchmark from the Grindavík school in 2023 and a second post-evacuation wave in 2026. It asks which aspects of well-being recovered, which persisted, and for whom.

### Previous research

Disasters that disrupt the lives and schooling of school-aged children and adolescents are associated with poorer well-being and more psychosomatic complaints (Kousky, 2016; La Greca et al., 2023; Osofsky et al., 2009; for a review see Auðardóttir & Gísladóttir, 2024). Similar consequences have followed other large disruptions of schooling, such as war and the COVID-19 pandemic (Bürgin et al., 2022; Betthäuser et al., 2023). These outcomes matter for adolescent health in their own right, and for students, well-being is also linked to learning (Bücker et al., 2018).

Displacement can disrupt several foundations of adolescents’ well-being at once: friendships, belonging to a school community, family routines, and a sense of safety. Recovery depends on how quickly these are rebuilt and on access to psychosocial services (Andrade et al., 2023; Abdi et al., 2023; Witt et al., 2024). Adolescents differ in their vulnerability and capacity to adapt, depending on individual, social, and contextual factors. Schools, families, and peers can provide protective systems that help them cope with displacement (Abdi et al., 2023; Masten & Narayan, 2012). Children’s responses to disasters are also associated with parental stress and coping, further adversities after the disaster, and the social support available to them (Pfefferbaum et al., 2015). This attention to disruption and adaptation informs our focus on life satisfaction, school belonging, psychosomatic complaints, substance use, and school absenteeism.

Post-disaster adjustment rarely follows a single trajectory. Longitudinal research distinguishes stable resilience, gradual recovery, chronic difficulties, and delayed-onset problems rather than a uniform return to pre-disaster well-being (Bonanno et al., 2024). In a review of eight studies that followed children after natural disasters, all eight identified groups whose posttraumatic stress symptoms remained low, declined, or remained elevated. Evidence for delayed onset was mixed (Lai et al., 2017). Two implications follow. First, a gap observed at one point after evacuation may later shrink, persist, or widen. Second, outcomes may not move together: global appraisals such as life satisfaction may rebound as new routines settle, while psychosomatic complaints or behavioural responses persist or emerge later. Recovery may also differ according to families’ socioeconomic status (see for example Lee and You, 2025) or between girls and boys. Disaster studies often report stronger emotional reactions among girls (Pfefferbaum et al., 2015), and in the 2025 wave, the gaps in life satisfaction and school belonging among Grindavík evacuees were larger for girls than for boys (Reimer et al., 2026). Less is known about whether such gender differences persist, narrow, or shift towards other outcomes as displacement continues. While potentially relevant, an analysis of different patterns by socioeconomic status are outside the scope of the current study because the data only includes a subjective measure of family status. This measure may be influenced not only by the displacement but also by how it influenced well-being.

Some of what follows an evacuation may reflect the student experience of moving as such rather than the disaster. Migration and residential mobility in childhood have been linked to emotional and behavioural difficulties (Andrade et al., 2023; Cotton, 2016). One reason for this is that mobility disrupts students’ friendship networks (South & Haynie, 2004), which are a major source of well-being among adolescents. Students entering new schools also face a higher risk of peer victimization (Tenhunen et al., 2024). Students who left Grindavík before the evacuation therefore offer a useful benchmark: difficulties shared with them may reflect mobility more generally, whereas difficulties found only among evacuees point more specifically to the evacuation. At the same time, prior movers are a differently selected group relative to evacuees; in many cases differences between movers and non-movers may be explained by other disadvantages, including the neighbourhoods they live in and selection processes leading to mobility, such as family instability (Cotton, 2016). In fact, previous research from Iceland on daily smoking among 15–16-year-olds found that adolescents who had moved to a new neighbourhood in the previous year were more likely to smoke daily than non-movers, even after adjusting for a range of family- and neighbourhood-level characteristics, including measures of social capital (Thorlindsson et al., 2012).

### Current study

This preregistered study uses the 2023, 2025, and 2026 waves of the Icelandic Youth Study to examine adolescent well-being after the Grindavík evacuation. We compare evacuees with a national reference group and with students who left Grindavík before November 2023, referred to as prior movers. The Grindavík school sample from 2023 provides a pre-evacuation benchmark. We address three research questions:

1. Do evacuees differ from the national reference group in life satisfaction, school belonging, psychosomatic symptoms, school absenteeism, and past-30-day substance use in 2025 and 2026? Did these gaps change between waves, and do they differ by gender?
2. Do life satisfaction, school belonging, and psychosomatic symptoms among evacuees in 2025 and 2026 depart from levels observed in the Grindavík school in 2023, after accounting for national time trends?
3. Do comparisons with prior movers suggest a well-being pattern distinctive to evacuation, or difficulties shared with students who left Grindavík earlier?

The analyses estimate adjusted differences between surveyed groups at each wave and changes in those differences between 2025 and 2026. Prior movers provide a mobility benchmark, and we use “evacuation-specific” to describe an observed pattern that distinguishes evacuees from prior movers. These comparisons do not identify the causal effects or individual adjustment trajectories. Continuing uncertainty about return provides context for interpreting the findings but is not directly measured.

## Method

### Data

We use repeated cross-sections from the Icelandic Youth Study (IYS), an annual nationwide school survey managed by the Educational Research Institute at the University of Iceland (Guðjohnsen et al., 2025). The Icelandic Youth Study was approved by the University of Iceland Science Ethics Committee for each yearly study. Parents/guardians were informed about the survey in advance and could withdraw their child from participation, and students were told that participation was voluntary, that they could skip any question or stop at any time, and that their answers were anonymous. Students answered electronically during school hours, and our analyses are limited to grades 6–10. The surveys were administered in March–May 2023, February–April 2025, and February–May 2026. School participation comprised 158 of 174 invited schools in 2023, 155 of 173 in 2025, and 153 schools in 2026. Each wave thus approximates a population survey of students in these grades. Published national student response rates ranged from approximately 68% to 79%, depending on wave and grade. Each wave is a separate cross-section.

In 2023, the Grindavík series consists of students attending the town’s only school, and the national reference group consists of students attending other schools. In 2025 and 2026, three questions establish whether students had ever lived in Grindavík, whether they still did, and whether those who had left moved before or after November 2023. We classify students as follows:

1. **Evacuees**: students who lived in Grindavík and relocated in November 2023 or later, as well as those who still reported residing in Grindavík.

2. **Prior movers**: students who lived in Grindavík but moved before November 2023.

3. **National reference group**: students who had never lived in Grindavík. Students whose responses do not permit classification are excluded.

The 2023 Grindavík school sample cannot be separated by their later displacement history. Prior movers are excluded from the 2025 and 2026 samples used in the pre-evacuation benchmark. Also, students who still reported a Grindavík address are counted as evacuees because everyday life and schooling in the town was not possible when the data was collected in spring 2025 and 2026. Finally, the survey does not collect data on when the prior moves actually occurred, but most seemed to be earlier than two school years before the evacuation (Reimer et al., 2026).

### Measures

Measures were similar across waves except where noted. In 2026, students received either the standard IYS or a Health Behaviour in School-aged Children (HBSC) questionnaire at random.

Life satisfaction uses a single ladder item from 0 (the worst possible life) to 10 (the best), with higher scores indicating greater life satisfaction (Cheung & Lucas, 2014).

School belonging is the mean of three items scored from 1 (strongly agree) to 4 (strongly disagree): “I feel like I belong at school,” “Other students seem to like me,” and “I feel lonely at school.” The first two are reversed so that higher scores indicate greater belonging; at least two items are required.

Psychosomatic symptoms are the mean of eight HBSC Symptom Checklist items covering complaints over the past six months: headache, stomach-ache, backache, dizziness, feeling low, irritability or bad temper, nervousness, and difficulties sleeping. Responses were harmonised to five frequency categories by combining rarely and never where necessary (for more details, see Supplementary Material).

School absenteeism is measured with two items on skipping a whole day or a class in the past two weeks. Responses range from 1 (never) to 4 (more than three times), and the models compare any school absence with none. In 2026, analyses are limited to the students receiving those questionnaire blocks.

Bullying victimisation was measured with six items adapted from the Olweus questionnaire, administered to grades 8–10 and referring to the past two months (Olweus, 1996).

Past-30-day substance use indicates any use of cigarettes, e-cigarettes, alcohol, or drunkenness. Valid HBSC responses take precedence, with IYS yes-or-no responses used otherwise. Students with no observed items are treated as missing. Absenteeism and substance use are analysed in 2025 and 2026 only.

### Analytical strategy

We compare evacuees with the national reference group separately in 2025 and 2026 and in pooled models with wave and evacuee × wave terms. Interactions estimate changes in the gap; linear combinations provide the 2026 contrasts. We re-estimate the 2025 comparisons reported previously (Reimer et al., 2026). The three well-being outcomes were preregistered, and absenteeism and substance use were added later. The study was preregistered at OSF (https://osf.io/h7jez). Departures from the preregistration are documented in Table S8 in the Supplementary Material.

We exclude careless respondents, respondents missing all three well-being outcomes, and those with unclassifiable Grindavík histories in 2025-2026. Models use complete cases, with sample sizes reported in the tables. Main models adjust for grade, gender, and origin-based non-Icelandic background and retain non-binary respondents. Gender comparisons use boys and girls only. As mentioned before, subjective socioeconomic status, a measure available in the survey, is omitted because evacuation may affect perceptions of family resources.

Continuous outcomes are estimated using linear models, with adjusted differences expressed in each model’s analytic-sample standard deviation units. Because standardisation is sample-specific, coefficient magnitudes are not directly comparable across models using different samples. Continuous models use school-clustered standard errors with the CR2 bias-reduction adjustment. Absenteeism and substance use are estimated using logistic models with school-clustered sandwich standard errors using HC2 adjustment. All confidence intervals are 95%.

Gender models include evacuee × wave × gender interactions. We report gender-specific group contrasts and direct tests of gender differences and changes in those differences. Supporting checks add gender interactions to the benchmark and prior-mover models, retaining their respective background controls.

For the three well-being outcomes, benchmark models include series, year, series × year interactions, grade, gender, and home language. The interactions estimate difference-in-differences contrasts: changes from the 2023 Grindavík–national gap to the corresponding gaps in 2025 and 2026, accounting for national time trends. We interpret these as descriptive departures from the pre-evacuation benchmark rather than causal effects, because a single pre-evacuation observation cannot establish parallel trends. Because the 2023 Grindavík estimate comes from a single school, we show it without a confidence interval. The confidence intervals for later departures treat this 2023 starting point as fixed and may therefore understate uncertainty. Adjusted means are averaged over the covariate distribution of each sample.

The prior-mover comparison uses a three-group × wave model for all five outcomes, with the same controls and uncertainty methods as the main comparison. We report group contrasts at each wave, changes in gaps, and direct evacuee–prior-mover contrasts.

Sensitivity analyses adjusting for home language repeat the main, gender, and prior-mover models for well-being and absenteeism, excluding students missing that item; in 2026, this restricts life satisfaction and symptoms to the block receiving it.

To assess sensitivity to changes in sample composition, we average predictions from the gender-interaction models using the same outcome-specific 2025 evacuee gender–grade distribution for both groups and waves. Within these cells, the distribution of non-Icelandic background is held at the 2025 national-reference distribution. Analyses are restricted to grades represented in all relevant groups and waves. This check does not address selective participation within gender–grade groups; its intervals condition on empirical weights, and absenteeism contrasts are marginal risk differences.

A further check restricts 2026 symptom models to IYS responses, retaining the full 2025 sample and corresponding main-model standard deviations. Additional cohort analyses compare expected grade cohorts across 2023–2026 and, more broadly, 2023–2025, adjusting for cohort, three-category gender, and home language. Full scoring and supplementary-analysis details are provided in the Supplementary Material.

## Results

### Sample characteristics

Table 1 describes the 2025 and 2026 samples by Grindavík category. The composition of both Grindavík groups shifted between waves, while the reference group remained stable. Among evacuees, the share of girls was lower in 2026 (36.7% vs. 42.3%) and the share with a non-Icelandic background higher (20.2% vs. 14.9%); among prior movers, the share of girls was higher (48.9% vs. 42.3%) and the mean grade lower (7.75 vs. 8.06). These differences likely reflect cohort turnover between waves and possibly selective relocation or nonresponse and are within the range expected from sampling variability at these group sizes. All models adjust for gender, grade, and background.

**Table 1.** Sample Characteristics by Group and Wave.

|  | 2025 |  |  | 2026 |  |  |
|---|---|---|---|---|---|---|
| Variable | Reference | Evacuees | Prior movers | Reference | Evacuees | Prior movers |
| n | 17,253 | 235 | 148 | 16,764 | 245 | 139 |
| Grade, M (SD) | 8.02 (1.41) | 8.04 (1.41) | 8.06 (1.41) | 8.02 (1.42) | 8.04 (1.42) | 7.75 (1.47) |
| Girls, % | 49.8 | 42.3 | 42.3 | 49.2 | 36.7 | 48.9 |
| Non-Icelandic background, % | 13.8 | 14.9 | 20.9 | 14.2 | 20.2 | 19.7 |
| Family well-off (1-5), M (SD) | 4.21 (0.75) | 4.27 (0.80) | 4.03 (0.83) | 4.24 (0.75) | 4.19 (0.95) | 4.05 (0.92) |
| Life satisfaction (0-10), M (SD) | 8.66 (1.85) | 8.27 (2.53) | 8.51 (2.19) | 8.75 (1.76) | 8.52 (2.46) | 8.31 (2.50) |
| School belonging (1-4), M (SD) | 3.12 (0.56) | 2.92 (0.62) | 3.05 (0.59) | 3.14 (0.56) | 3.01 (0.66) | 2.98 (0.73) |
| Psychosomatic symptoms (1-5), M (SD) | 2.30 (1.01) | 2.54 (1.13) | 2.64 (1.12) | 2.29 (0.97) | 2.56 (1.10) | 2.55 (1.06) |
| Any school absence, % | 25.6 | 37.4 | 40.8 | 22.5 | 36.0 | 32.6 |
| Any past-30-day substance use, % | 6.6 | 11.6 | 16.9 | 6.2 | 15.1 | 10.1 |

*Note.* n denotes the group sample after screening. Means and percentages use available responses for each variable; denominators therefore vary by row. Means are on the original measurement scales.

### Evacuee–national differences in 2025 and 2026

In 2025, evacuees reported lower life satisfaction (*B* = -0.21, 95% CI [-0.39, -0.04]), and school belonging (*B* = -0.37, 95% CI [-0.52, -0.23]) than the national reference group (Table 2). They also reported more psychosomatic symptoms (*B* = 0.26, 95% CI [0.07, 0.45]), and higher odds for school absence (OR = 1.69, 95% CI [1.22, 2.32]) and any past-30-day substance use (OR = 1.87, 95% CI [1.18, 2.96]).

**Table 2.** Evacuee Contrasts with the National Reference Group in 2025 and 2026.

| Contrast | Life satisfaction | School belonging | Psychosomatic symptoms | Absenteeism (OR) | Substance use (OR) |
|---|---|---|---|---|---|
| 2025 | **-0.21 [-0.39, -0.04]** | **-0.37 [-0.52, -0.23]** | **0.26 [0.07, 0.45]** | **1.69 [1.22, 2.32]** | **1.87 [1.18, 2.96]** |
| 2026 | -0.11 [-0.28, 0.07] | -0.21 [-0.46, 0.04] | **0.29 [0.08, 0.49]** | **1.81 [1.26, 2.60]** | **2.59 [1.75, 3.84]** |
| Change, 2025 to 26 | 0.09 [-0.13, 0.30] | 0.14 [-0.14, 0.42] | 0.03 [-0.17, 0.24] | 1.07 [0.73, 1.56] | 1.42 [0.82, 2.44] |

*Note.* Estimates compare evacuees with the national reference, adjusting for grade, gender, and non-Icelandic background; prior movers are excluded. Continuous outcomes use wave-specific standardisation; binary outcomes are odds ratios. Changes come from pooled models, with pooled standardisation, rather than subtraction of yearly estimates. Brackets show 95% school-clustered CIs (CR2 continuous; HC2 binary); bold indicates exclusion of 0 or 1, respectively. Questionnaire rotation restricts 2026 belonging and absenteeism samples. Analytic n (2025/2026): life satisfaction, 17,254/16,688; belonging, 17,368/11,712; symptoms, 17,310/16,733; absenteeism, 17,423/11,730; substance use, 17,392/16,783.

In 2026, symptoms remained higher than in the national reference group (*B* = 0.29, 95% CI [0.08, 0.49]), as did the odds of school absence (OR = 1.81, 95% CI [1.26, 2.60]) and substance use (OR = 2.59, 95% CI [1.75, 3.84]). Life satisfaction (B = -0.11, 95% CI [-0.28, 0.07]) and belonging (B = -0.21, 95% CI [-0.46, 0.04]) were directionally lower, with both intervals including zero.

The pooled estimates were compatible with no change in any of the five gaps between 2025 and 2026: all continuous-outcome change intervals included zero, and both odds-ratio intervals included 1 (Table 2).

### Gender differences in the evacuee–national reference gaps

In 2025, the evacuee–national gaps were larger for girls than boys on life satisfaction (girl-minus-boy difference: *B* = −0.60, 95% CI [−1.00, −0.19]) and school belonging (*B* = −0.39, 95% CI [−0.72, −0.07]). These gender differences narrowed by 2026: the changes were *B* = 0.62, 95% CI [0.10, 1.14], and *B* = 0.50, 95% CI [0.08, 0.92], respectively. In 2026, the life-satisfaction and belonging gaps for both genders had intervals including zero (Table 3).

**Table 3.** Evacuee Contrasts by Gender in 2025 and 2026.

| Contrast | Life satisfaction | School belonging | Psychosomatic symptoms | Absenteeism (OR) | Substance use (OR) |
|---|---|---|---|---|---|
| Boys, 2025 | 0.02 [-0.26, 0.30] | **-0.22 [-0.39, -0.05]** | **0.30 [0.02, 0.59]** | **1.89 [1.32, 2.70]** | 1.75 [0.83, 3.70] |
| Girls, 2025 | **-0.58 [-0.83, -0.33]** | **-0.62 [-0.88, -0.35]** | **0.25 [0.03, 0.47]** | 1.36 [0.80, 2.33] | 1.57 [0.82, 3.01] |
| Boys, 2026 | -0.08 [-0.31, 0.15] | -0.23 [-0.54, 0.07] | **0.44 [0.15, 0.73]** | **2.07 [1.33, 3.22]** | **2.75 [1.67, 4.55]** |
| Girls, 2026 | -0.06 [-0.36, 0.24] | -0.13 [-0.48, 0.22] | -0.00 [-0.26, 0.25] | 1.31 [0.64, 2.71] | 1.79 [0.78, 4.14] |

*Note.* Estimates compare evacuees with national peers of the same gender, adjusting for grade and non-Icelandic background. Continuous outcomes are standardised differences; binary outcomes are odds ratios. Brackets show 95% school-clustered CIs (CR2 continuous; HC2 binary); bold indicates exclusion of 0 or 1, respectively. Models include boys and girls only and exclude prior movers. Questionnaire rotation restricts 2026 belonging and absenteeism samples. Analytic n: life satisfaction, 33,110; belonging, 28,320; symptoms, 33,205; absenteeism, 28,389; substance use, 33,326. Some evacuee gender cells have fewer than 10 events.

The 2025 symptom-gap estimates were similar in size, and the direct gender-difference interval included zero. Between waves, the gap shifted towards boys relative to girls (girl-minus-boy change: B = −0.391, 95% CI [−0.780, −0.002]), with the upper limit close to zero. In 2026, boys’ symptoms remained higher than those of their national peers (B = 0.44, 95% CI [0.15, 0.73]); girls’ estimate was near zero, with an interval including zero. Boys’ own change interval included zero.

Boys had higher odds of school absence than their national peers in 2025 (OR = 1.89, 95% CI [1.32, 2.70]) and 2026 (OR = 2.07, 95% CI [1.33, 3.22]), girls’ intervals included 1. The direct 2025 gender contrast (OR = 0.72, 95% CI [0.44, 1.19]) did not establish a gender difference. Neither the gender difference nor boys’ gap clearly changed between waves.

Boys had higher odds of substance use in 2026 (OR = 2.75, 95% CI [1.67, 4.55]). The other three gender-specific intervals included 1. The direct 2025 gender contrast and its change between waves also had intervals including 1, as did boys’ own change. Some gender cells have fewer than 10 events.

### Departures from the pre-evacuation benchmark

In 2023, point differences between the Grindavík school and the national reference group were small: life satisfaction *B* = 0.03, belonging *B* = -0.06, and symptoms *B* = -0.05 (Figure 1, Table S1). Because the series represents one school, school-clustered intervals cannot describe its precision. Later departure intervals also use this single-school starting point.

**Figure 1.**
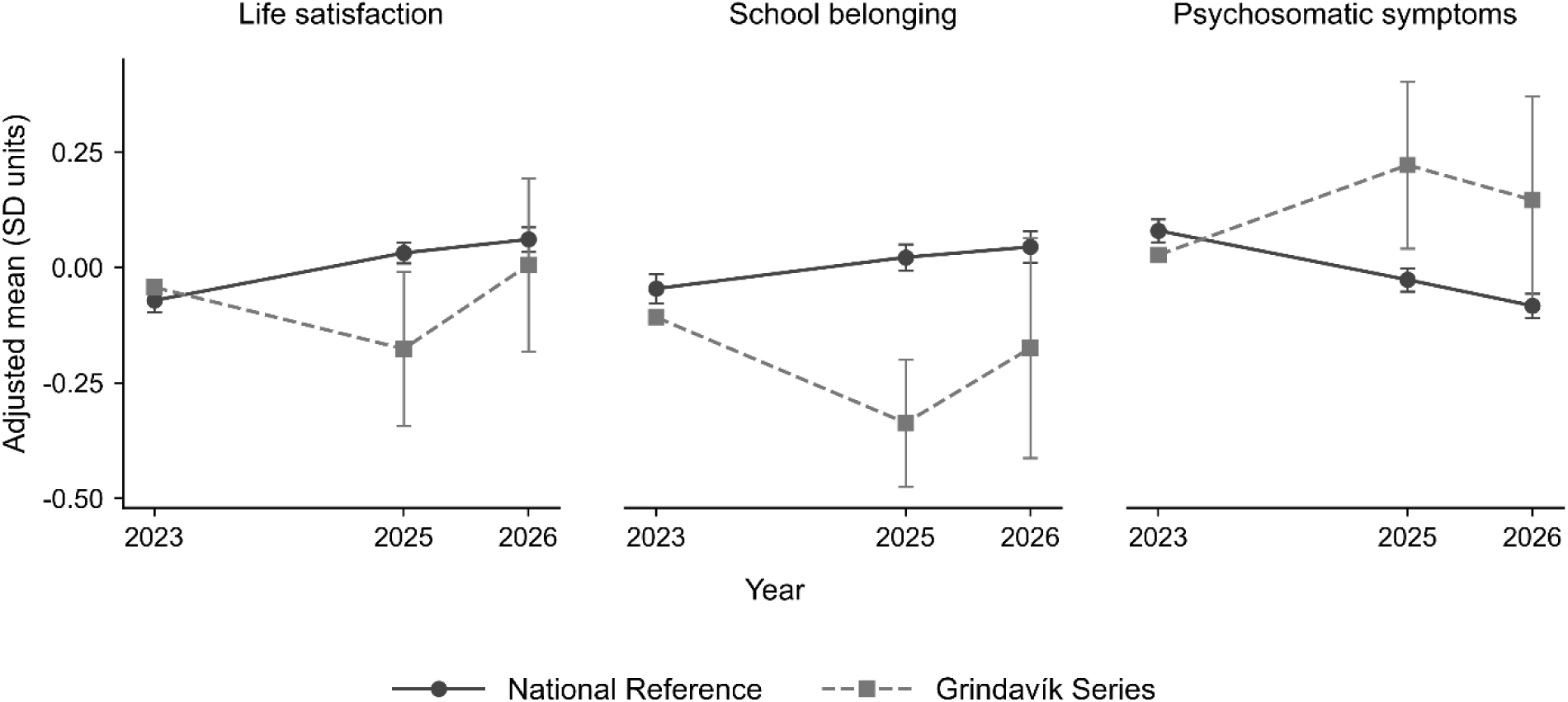
Adjusted Means for the Grindavík Series and the National Reference Group. Note. Means are adjusted predictions in benchmark-sample SD units, averaged over grade, gender, and home language. Bars show 95% school-clustered CR2 CIs. The 2023 Grindavík point represents one school, includes later prior movers, and is shown without a CI. Missing home-language responses are excluded. Questionnaire rotation restricts 2026 life satisfaction and symptoms to the home-language block and belonging to eligible grades.

After national time trends, the 2025 departures from the 2023 gaps were toward lower life satisfaction (*B* = -0.24, 95% CI [-0.41, -0.06]), and belonging (*B* = -0.30, 95% CI [-0.44, - 0.15]), and higher symptoms (*B* = 0.30, 95% CI [0.11, 0.49]). The symptom departure persisted in 2026 (*B* = 0.28, 95% CI [0.05, 0.51]). Life satisfaction (*B* = -0.08, 95% CI [-0.28, 0.11]), and belonging (*B* = -0.16, 95% CI [-0.41, 0.09]) were directionally lower relative to the 2023 gaps, with both intervals including zero. In 2026, the life-satisfaction and symptom estimates are limited to students receiving the home-language item, and the belonging estimates to those receiving the belonging items.

### Gender differences relative to the 2023 benchmark

Girls’ 2023 point contrasts with national peers were small *B* = 0.07 for life satisfaction and *B* = -0.00 for belonging. From 2023 to 2025, the girl-minus-boy difference in the Grindavik-national gap shifted towards lower life satisfaction (*B* = -0.66, 95% CI [-1.05, -0.27]) and belonging (*B* = -0.48, 95% CI [-0.80, -0.17]) using the same single-school starting point (Table S2). In 2026, gender difference point estimates were small, with wide intervals including zero: life satisfaction (*B* = -0.02, 95% CI [-0.42, 0.38]), belonging (*B* = 0.09, 95% CI [-0.29, 0.48]). These estimates use the benchmark sample, its standard deviation, and the home-language adjustment, so their magnitudes are not directly comparable with the main 2025–2026 gender contrasts.

### Prior movers as a mobility benchmark

In 2025, prior movers reported more psychosomatic symptoms (B = 0.38, 95% CI [0.20, 0.56]), and higher odds of school absence (OR = 1.88, 95% CI [1.29, 2.74]) and substance use (OR = 2.92, 95% CI [1.84, 4.64]) than the national reference group, as did evacuees (Table 4). Direct evacuee–prior-mover contrasts on these outcomes had intervals including zero or 1.

**Table 4.** Evacuee and Prior-Mover in Comparison to National Reference in 2025 and 2026.

| Contrast | Life satisfaction | School belonging | Psychosomatic symptoms | Absenteeism (OR) | Substance use (OR) |
|---|---|---|---|---|---|
| Evacuees, 2025 | **-0.21 [-0.39, -0.03]** | **-0.36 [-0.51, -0.22]** | **0.26 [0.07, 0.45]** | **1.69 [1.22, 2.32]** | **1.86 [1.18, 2.94]** |
| Prior movers, 2025 | -0.09 [-0.28, 0.10] | -0.11 [-0.31, 0.09] | **0.38 [0.20, 0.56]** | **1.88 [1.29, 2.74]** | **2.92 [1.84, 4.64]** |
| Evacuees versus prior movers, 2025 | -0.12 [-0.39, 0.15] | **-0.25 [-0.49, -0.01]** | -0.12 [-0.36, 0.12] | 0.90 [0.59, 1.36] | 0.64 [0.34, 1.18] |
| Evacuees, 2026 | -0.12 [-0.30, 0.05] | -0.23 [-0.48, 0.03] | **0.29 [0.09, 0.50]** | **1.80 [1.26, 2.59]** | **2.64 [1.79, 3.88]** |
| Prior movers, 2026 | -0.22 [-0.48, 0.04] | -0.28 [-0.58, 0.03] | **0.28 [0.10, 0.46]** | **1.66 [1.07, 2.58]** | **1.87 [1.07, 3.27]** |
| Evacuees versus prior movers, 2026 | 0.10 [-0.20, 0.40] | 0.05 [-0.34, 0.44] | 0.01 [-0.24, 0.27] | 1.09 [0.62, 1.90] | 1.41 [0.76, 2.64] |
| Change for evacuees | 0.09 [-0.13, 0.30] | 0.14 [-0.14, 0.42] | 0.03 [-0.17, 0.24] | 1.07 [0.73, 1.56] | 1.42 [0.82, 2.44] |
| Change for prior movers | -0.13 [-0.41, 0.15] | -0.16 [-0.47, 0.14] | -0.10 [-0.32, 0.12] | 0.88 [0.51, 1.52] | 0.64 [0.35, 1.15] |

*Note.* Estimates compare each group with the national reference, adjusting for grade, gender, and non-Icelandic background. Yearly contrasts and changes come from pooled models. Continuous outcomes are standardised within the pooled three-group sample; binary outcomes are odds ratios. Brackets show 95% school-clustered CIs (CR2 continuous; HC2 binary); bold indicates exclusion of 0 or 1, respectively. Contrasts are against the national reference unless labelled otherwise. Analytic n: life satisfaction, 34,224; belonging, 29,313; symptoms, 34,326; absenteeism, 29,386; substance use, 34,460.

Prior movers were directionally lower than national peers on life satisfaction and belonging, with both intervals including zero. Evacuees were lower than prior movers on belonging (B = -0.25, 95% CI [-0.49, -0.01]), with the upper limit just below zero. The direct life-satisfaction contrast was also negative but uncertain (B = -0.12, 95% CI [-0.39, 0.15]). In 2026, both groups continued to report more symptoms and higher odds of absence and substance use than national peers. All direct evacuee-prior mover contrasts that year included zero or 1. Pooled change estimates were compatible with no change in any outcome gap for either group.

### Gender differences among prior movers

In 2025, prior-mover boys and girls were directionally lower than national peers on life satisfaction and belonging, with all four intervals including zero; girls’ estimates were close to zero (Table S2). Evacuee girls were about half a standard deviation below prior-mover girls on both life satisfaction (B = -0.49, 95% CI [-0.92, -0.06]) and belonging (B = -0.58, 95% CI [-1.01, -0.15]), with intervals of similar width. For boys, both direct comparison intervals included zero.

In 2026, prior-mover girls were lower than national peers on belonging (B = -0.48, 95% CI [-0.92, -0.04]), with a wide interval. Evacuee girls were directionally lower (B = -0.13, 95% CI [-0.48, 0.22]), with an interval including zero; the direct comparison between the two groups also included zero. The distinctive observed evacuee-girl pattern concerned 2025.

Prior-mover boys and girls had higher odds of substance use than national peers in 2025 (OR = 2.70, 95% CI [1.36, 5.39], and OR = 3.43, 95% CI [1.76, 6.67], respectively). Their 2026 odds ratios remained above 1, with wide intervals including 1. Direct evacuee–prior-mover intervals included 1 for both genders in both years; some gender cells had fewer than 10 events.

### Sensitivity analyses

Sensitivity analyses adjusting for home language retained the 2025 evacuee–national pattern. In 2026, symptoms remained directionally higher, but with an interval including zero on the other sample. Other outcomes retained the main patterns (Table S4).

Holding the 2025 evacuee gender–grade composition constant produced similar estimates to using each wave’s observed composition. For this target, all change intervals for the three well-being outcomes and absenteeism included zero, including life satisfaction (B = 0.17, 95% CI [−0.03, 0.36]).

Restricting 2026 to IYS responses yielded a smaller positive symptom estimate with greater uncertainty (B = 0.23, 95% CI [−0.01, 0.47]); the change interval also included zero (B = −0.02, 95% CI [−0.27, 0.23]; Table S5). This restriction also changes sample composition and does not isolate a response-scale effect.

The cohort-progression analyses showed that all three-wave cohort departure intervals included zero. The broader 2023–2025 comparison showed lower belonging relative to the pre-evacuation gap; life-satisfaction and symptom intervals included zero (Table S7).

## Discussion

This study examined adolescent well-being after the Grindavík evacuation in two waves, both collected while the town was still rated high risk and return remained unresolved. Before the evacuation, students at the Grindavík school reported well-being close to national levels. In 2025, after national trends, evacuees’ life satisfaction and school belonging were lower and their psychosomatic symptoms higher than this benchmark would suggest, and they reported more school absence and substance use than national peers. In 2026, symptoms, absence, and substance use remained elevated, whereas the life-satisfaction and belonging gaps were smaller, with intervals including zero. As the change estimates did not establish narrowing, this pattern indicates continuing difficulties rather than recovery.

The outcomes did not move together. Between 2025 and 2026, the life-satisfaction and belonging gaps moved towards national levels, whereas the symptom gap remained of similar size relative to both national peers and the 2023 benchmark. Although the change estimates did not establish narrowing, this pattern is consistent with asynchronous, domain-specific recovery, in which dimensions of well-being follow different trajectories after a disaster (Bonanno et al., 2024). Life satisfaction and school belonging are evaluative and relational: they can recover as adolescents form new friendships, settle into host schools, and recalibrate their standards to changed circumstances (Lucas, 2007). Psychosomatic complaints, which combine bodily symptoms such as headache and sleep difficulties with nervousness and low mood, are better understood as expressions of sustained stress and can persist for as long as the stressors do (McEwen, 1998). After the 2010 Eyjafjallajökull eruption, parents in exposed areas were more likely than parents in the comparison area to report anxiety or worries in their children. Among exposed children, the study detected no significant decline in parent-reported symptoms between 2010 and 2013 (Hlodversdottir et al., 2018). In 2026, those stressors had not ended: families had moved repeatedly, many still regarded their housing as temporary (Forsætisráðuneytið, 2025), and the question of return remained open. One reading of our findings is therefore that adolescents had largely re-established their social and school lives, whereas the strain of prolonged displacement and uncertainty remained present in body and mind. Alternatively, symptoms may simply recover more slowly or reflect delayed responses; separating these explanations requires longitudinal data. Either way, improvement in global well-being should not be read as a sign that the consequences of displacement have passed.

These gaps emerged under comparatively favourable conditions. The Icelandic response combined financial support for displaced families, continued schooling, and psychosocial services within a well-resourced Nordic welfare state. In many respects, the Grindavík evacuation thus approximates a best-case scenario for displaced adolescents. That symptoms, school absence, and substance use nevertheless remained elevated more than two years later suggests that material security and institutional continuity, although necessary, do not by themselves protect adolescents from the strain of prolonged displacement. Where support is weaker, the consequences are likely to be larger.

### Evacuation and other residential mobility

Evacuees and prior movers both reported more psychosomatic symptoms, school absence, and substance use than national peers in both years, and direct comparisons did not distinguish the two groups on these outcomes. Although shared outcomes need not reflect shared processes, this similarity suggests that part of what follows an evacuation reflects relocation and school change more generally, consistent with research linking mobility to weaker school connectedness (Capp & Sullivan, 2020) and, in Iceland, to adolescent daily smoking (Thorlindsson et al., 2012). Eighteen months after the Fort McMurray wildfire, students reported more tobacco use than students surveyed three years earlier in Red Deer, although the tobacco questions covered the past month in Fort McMurray and the past year in Red Deer. Brown et al. (2019) found no significant difference in alcohol use. In Grindavík, our combined measure leaves open which substances accounted for the higher odds of use among evacuees and prior movers.

As previously reported for 2025 (Reimer et al., 2026), the clearest evacuation-specific contrast concerned school belonging, particularly among girls; the estimate was imprecise, and by 2026 the contrast was no longer evident. Belonging is also where uncertainty about return might matter most. Unlike most moves, the evacuation was involuntary and open-ended, so investing in a new school community may have seemed less worthwhile while ties to the former one persisted. As uncertainty was not measured, this remains a hypothesis. Practically, support should reach residentially mobile students whatever their reason for moving (Tenhunen et al., 2024), while evacuated students may additionally need help rebuilding a sense of belonging, a task in which schools are central (Smawfield, 2013).

### Gender differences across outcomes and time

Gender patterns differed across outcomes and changed over time. In 2025, the life-satisfaction and belonging gaps were larger for evacuee girls than boys, a pattern not visible at the 2023 benchmark, and evacuee girls were about half a standard deviation below prior-mover girls on both outcomes. By 2026, these gender differences had narrowed, and girls’ symptom gap was no longer evident, whereas boys’ symptom and absence gaps persisted in both years. Direct tests did not establish gender differences in absence or substance use. Because the narrowing coincided with a fall in girls’ share of the evacuee sample, it may partly reflect who participated rather than improved well-being. The findings therefore argue against a general account of greater vulnerability among girls and for attention to outcomes and timing.

## Limitations

Because the data are repeated cross-sections, they cannot trace individual trajectories, and changes in group gaps may reflect changes in well-being, group composition, or participation. Composition did shift between waves: girls’ share of the evacuee sample fell from 42.3% to 36.7%, particularly in grades 9–10, while the 2026 prior movers included more girls and were younger on average. Holding the 2025 evacuee gender–grade composition constant had little effect on the estimates, but neither this check nor covariate adjustment addresses selective participation within groups. If girls with poorer well-being were less likely to participate in 2026, their gaps would be underestimated; whether this occurred is unknown.

The design is observational. It cannot formally establish that the gaps were caused by the evacuation, separate the contributions of different housing and schooling arrangements, or identify disrupted school connections as the mechanism behind lower belonging. Uncertainty about return, central to our interpretation, was not measured directly, and we do not know how students’ expectations of return differed between individuals or changed between waves.

The pre-evacuation benchmark rests on a single school at one time point and cannot show whether Grindavík would otherwise have followed national trends. Its 2023 sample includes students later classified as prior movers, and differences in analytic-sample standard deviations mean that benchmark and 2025–2026 coefficients are not directly comparable in magnitude. The benchmark also does not cover school absence or substance use, so we cannot establish when those gaps emerged.

Questionnaire rotation restricted some analytic samples, and complete-case analysis may introduce selection if missing responses are related to well-being. Harmonising symptom response categories does not establish measurement equivalence. The home-language and IYS-only checks yielded smaller, less precise symptom estimates; because they also change sample composition, they cannot isolate response-scale effects.

Despite these limitations, the design makes several alternative explanations unlikely. Because the whole town was evacuated, evacuees were not selected into displacement by family circumstances, as residential movers typically are. The 2023 benchmark shows that Grindavík students did not stand out from their national peers before the evacuation, even after several years of seismic unrest, and the 2025 departures from that benchmark were estimated net of national time trends. Pre-existing differences, national developments, or self-selection are therefore unlikely to account for the post-evacuation gaps. Although the data cannot follow individuals over time, we consider the overall pattern difficult to reconcile with explanations unrelated to the displacement.

## Conclusions

Natural disasters are a growing source of uncertainty in children’s lives. Climate change is increasing the frequency and intensity of many weather-related hazards, and floods, storms, droughts, and wildfires already account for tens of millions of child displacements worldwide (IPCC, 2023; UNICEF, 2023). Although the Grindavík crisis was geological, it shares a defining feature of many such displacements: an open-ended period of uncertainty about return. This uncertainty concerns not only whether return will become safe, but also, as displacement lengthens and children settle into new schools and friendships, whether families will still want to go back. As such events become more common, including in high-income countries, understanding how adolescents fare during this period becomes increasingly relevant for public health.

Our findings show that difficulties among displaced adolescents can outlast the immediate aftermath of an evacuation, even under close to best-case conditions of material support, schooling, and social services. More than two years after leaving Grindavík, evacuees still reported more psychosomatic symptoms, school absence, and substance use than national peers. These difficulties were shared with students who had moved for other reasons, whereas the main evacuation-specific contrast concerned school belonging. Both waves captured a period in which return remained unresolved.

Since our data were collected, the formal barrier to return has been lifted: in summer 2026, Civil Protection lowered the town’s risk level (Almannavarnir, 2026), and the Grindavík school reopened in August. Yet only about 30 students enrolled, and most families remain elsewhere (Víkurfréttir, 2026). After almost three years away, return is no longer the default. The question has shifted from whether families can go back to whether they want to, and for many the answer appears to be no, or not yet. For most adolescents, displacement is thus turning into permanent relocation, while for families who remain undecided, uncertainty continues in a new form. Future waves can show whether gaps in belonging and well-being diminish as these situations settle. Support for displaced adolescents should address not only material needs, symptoms, and continuity of schooling, but also friendships, attachment to school communities, and a sense of belonging as outcomes of recovery in their own right.

## Acknowledgements

The authors would like to thank all the schools and students who participated in the data collection for the Icelandic Youth Study and the survey team at the University of Iceland. We also extend our special thanks to the Grindavík committee members who provided helpful background information for this article.

## Statements and Declarations

### Data availability statement

The data are not publicly available but can be requested from the Educational Research Institute at the University of Iceland. The analytic code is available at repository: [DOI HERE].

### Author contributions statement

David Reimer: Conceptualization, Writing – original draft, Writing – review & editing. Juuso Repo: Conceptualization, Formal analysis, Writing – original draft, Writing – review & editing. Elina Kilpi-Jakonen: Writing – review & editing. Kolbrún Þ. Pálsdóttir: Writing – review & editing. All authors approved the final version of the manuscript and agree to be accountable for all aspects of the work.

