## Supplementary Material for "Displaced by a Volcano: Adolescent Well-Being in Uncertain Times in Iceland, 2023–2026"

Online Supplementary Material for:
"Displaced by a Volcano: Adolescent Well-Being in Uncertain Times in Iceland, 2023–2026"

**Table S1**

*Grindavík Series Contrasts After the National Time Trend*

| Contrast | Life satisfaction | School belonging | Psychosomatic symptoms |
|---|---|---|---|
| 2023 | 0.03 | -0.06 | -0.05 |
| Departure in 2025 | -0.24 [-0.41, -0.06] | -0.30 [-0.44, -0.15] | 0.30 [0.11, 0.49] |
| Departure in 2026 | -0.08 [-0.28, 0.11] | -0.16 [-0.41, 0.09] | 0.28 [0.05, 0.51] |

*Note.* Estimates are standardised differences in the pooled 2023–2026 sample, adjusted for grade, gender, and home language. The 2023 row shows the Grindavík–national gap; later rows show departures from that gap after national trends. Brackets show 95% school-clustered CR2 CIs. The single-school 2023 point has no CI; later intervals treat it as fixed and may understate uncertainty. Later prior movers are included in 2023 but excluded thereafter. Missing home-language responses are excluded; questionnaire rotation restricts 2026 samples. Analytic n: life satisfaction, 45,726; belonging, 46,038; symptoms, 45,876.

**Table S2**

*Life Satisfaction and School Belonging in the Gender Checks*

| Contrast | Life satisfaction | School belonging |
|---|---|---|
| 2023 school, girls | 0.07 | -0.00 |
| 2023 school, boys | -0.01 | -0.07 |
| Change in the gender difference, 2023 to 2025 | **-0.66 [-1.05, -0.27]** | **-0.48 [-0.80, -0.17]** |
| Gender difference, 2026 | -0.02 [-0.42, 0.38] | 0.09 [-0.29, 0.48] |
| Prior-mover girls, 2025 | -0.09 [-0.41, 0.23] | -0.03 [-0.34, 0.27] |
| Prior-mover boys, 2025 | -0.05 [-0.30, 0.20] | -0.17 [-0.42, 0.08] |
| Evacuee girls versus prior-mover girls, 2025 | **-0.49 [-0.92, -0.06]** | **-0.58 [-1.01, -0.15]** |
| Evacuee boys versus prior-mover boys, 2025 | 0.07 [-0.34, 0.48] | -0.05 [-0.32, 0.21] |
| Prior-mover girls, 2026 | -0.31 [-0.65, 0.04] | **-0.48 [-0.92, -0.04]** |
| Evacuee girls, 2026 | -0.06 [-0.36, 0.24] | -0.13 [-0.48, 0.22] |

*Note.* Estimates are adjusted standardised differences; brackets show 95% school-clustered CR2 CIs. Benchmark rows use the 2023–2026 sample SD and home-language adjustment; prior-mover rows use the 2025–2026 three-group SD and origin-based adjustment, so magnitudes are not directly comparable across blocks. Contrasts are against national peers unless labelled otherwise. Gender differences are girl minus boy; change is 2025 minus 2023, with negative values indicating a shift towards lower scores for girls. The single-school 2023 points include later prior movers and have no CIs. Benchmark analyses exclude missing home language, restricting 2026 life satisfaction to that questionnaire block. Analytic n (life satisfaction/belonging): benchmark, 44,369/44,657; prior-mover check, 33,383/28,544.

**Table S3**

*Substance-Use Contrasts by Gender in the Three-Group Model*

| Gender | Contrast | 2025 | 2026 |
|---|---|---|---|
| Boys | Evacuees versus national | 1.75 [0.83, 3.70] | **2.76 [1.67, 4.55]** |
| Boys | Prior movers versus national | **2.70 [1.36, 5.39]** | 1.47 [0.56, 3.81] |
| Boys | Evacuees versus prior movers | 0.65 [0.24, 1.76] | 1.88 [0.65, 5.45] |
| Girls | Evacuees versus national | 1.57 [0.82, 3.01] | 1.79 [0.78, 4.14] |
| Girls | Prior movers versus national | **3.43 [1.76, 6.67]** | 1.62 [0.66, 4.02] |
| Girls | Evacuees versus prior movers | 0.46 [0.19, 1.11] | 1.10 [0.30, 4.12] |

*Note.* Odds ratios indicate any past-30-day substance use; brackets show 95% school-clustered HC2 CIs. The pooled group × wave × gender model adjusts for grade and non-Icelandic background and includes boys and girls only. Some gender cells have fewer than 10 events. Analytic n = 33,601.

Home-language adjustment retained the 2025 evacuee–national patterns. In 2026, symptoms remained directionally higher, with an interval including zero; the other outcomes retained the main patterns (Table S4). Evacuee girls remained below national peers in 2025 on life satisfaction (B = -0.57, 95% CI [-0.81, -0.33]) and belonging (B = -0.63, 95% CI [-0.89, -0.36]). The 2026 symptom gaps remained positive for evacuee boys (B = 0.32, 95% CI [0.02, 0.62]) and prior movers (B = 0.37, 95% CI [0.11, 0.63]). The 2025 evacuee–prior-mover belonging contrast was directionally negative, with an interval including zero (B = -0.24, 95% CI [-0.48, 0.00]).

**Table S4**

*Evacuee Contrasts with the Home-Language Item*

| Contrast | Life satisfaction | School belonging | Psychosomatic symptoms | Absenteeism (OR) |
|---|---|---|---|---|
| 2025 | -0.22 [-0.39, -0.04] | -0.37 [-0.51, -0.22] | 0.25 [0.06, 0.44] | 1.66 [1.21, 2.27] |
| 2026 | -0.04 [-0.24, 0.16] | -0.20 [-0.45, 0.04] | 0.22 [-0.01, 0.45] | 1.80 [1.26, 2.58] |
| Change | 0.16 [-0.06, 0.38] | 0.14 [-0.14, 0.42] | -0.02 [-0.27, 0.22] | 1.08 [0.73, 1.59] |

*Note.* Models adjust for grade, gender, and home language. Continuous outcomes use wave-specific standardisation; absenteeism estimates are odds ratios. Changes come from pooled models, with pooled standardisation, rather than subtraction of yearly estimates. Brackets show 95% school-clustered CIs (CR2 continuous; HC2 absenteeism). Missing home-language responses are excluded, restricting 2026 life satisfaction and symptoms to that questionnaire block. Analytic n (2025/2026): life satisfaction, 17,290/11,621; belonging, 17,404/11,736; symptoms, 17,346/11,667; absenteeism, 17,460/11,755.

### IYS-only symptom sensitivity

Table S5 compares the combined-version and IYS-only symptom estimates, retaining the full 2025 sample and main-model standard deviations. The restricted-sample 2026 and change intervals included zero. Excluding HBSC responses also changes sample composition, so this comparison does not isolate a response-scale effect.

**Table S5**
*Psychosomatic Symptom Contrasts with 2026 Restricted to IYS Responses*

| Contrast | Combined versions, B [95% CI] | IYS-only sensitivity, B [95% CI] |
|---|---|---|
| 2025, wave-specific | **0.260 [0.069, 0.451]** | **0.260 [0.069, 0.451]** |
| 2026, wave-specific | **0.289 [0.084, 0.494]** | 0.230 [-0.010, 0.470] |
| Pooled model: 2025 contrast | **0.260 [0.067, 0.453]** | **0.261 [0.068, 0.454]** |
| Pooled model: 2026 contrast | **0.295 [0.090, 0.500]** | 0.237 [-0.001, 0.475] |
| Pooled model: change in the gap | 0.035 [-0.171, 0.240] | -0.024 [-0.273, 0.226] |

*Note.* Positive contrasts indicate more frequent symptoms among evacuees than the national reference group. Models adjust for grade, gender, and origin-based non-Icelandic background, with CR2 standard errors clustered at school. The sensitivity analysis retains the full 2025 sample and restricts 2026 to IYS responses. The 2026 model includes 157 evacuees and 11,486 national-reference respondents, compared with 234 and 16,499, respectively, in the combined-version model. Standardisation retains the corresponding main-model standard deviations. HBSC responses occur only in grades 6, 8, and 10, so excluding them also changes sample composition; this comparison does not isolate a response-scale effect.

**Table S6**
*Bullying Victimisation Contrasts with the National Reference Group*

| Contrast | B [95% CI] | Analytic n | Schools |
|---|---|---|---|
| 2025 | -0.02 [-0.20, 0.17] | 10,245 | 130 |
| 2026 | 0.01 [-0.21, 0.24] | 6,622 | 127 |
| Change, 2025–2026 | 0.04 [-0.26, 0.33] | 16,867 | 134 |

*Note.* Higher scores indicate more frequent victimisation in the past two months (six-item mean; ≥3 answers required). Estimates use wave-specific standardisation; change is the pooled evacuee × wave interaction, with pooled standardisation, rather than subtraction of yearly estimates. Brackets show 95% school-clustered CR2 CIs. Models adjust for grade, gender, and non-Icelandic background, retaining non-binary respondents and excluding prior movers. Careless responses and respondents missing all three well-being outcomes are excluded. Complete-case n (evacuees/national reference): 132/10,113 in 2025; 86/6,536 in 2026.

The non-preregistered bullying contrasts were close to zero in both years, with all wave-specific and change intervals including zero (Table S6). These estimates do not establish equivalence.

### Cohort-progression models

Cohort-progression models compare repeated cross-sections of expected grade cohorts. Grades 6–7 in 2023 progress to grades 8–9 in 2025 and grades 9–10 in 2026; the broader two-wave comparison additionally includes grade 8 in 2023 and grade 10 in 2025. Both models retain three-category gender adjustment. All three-wave departure intervals included zero. The broader 2023–2025 comparison showed lower belonging relative to the pre-evacuation gap, while life-satisfaction and symptom intervals included zero (Table S7).

**Table S7**

*Departures from the 2023 Grindavik-National Gap in the Cohort-Progression Analyses*

| Outcome | Three-wave model: 2025 | Three-wave model: 2026 | Two-wave model: 2025 |
|---|---|---|---|
| Life satisfaction | -0.23 [-0.51, 0.06] | 0.14 [-0.15, 0.44] | -0.18 [-0.41, 0.04] |
| School belonging | -0.16 [-0.42, 0.10] | 0.01 [-0.31, 0.33] | -0.27 [-0.50, -0.04] |
| Psychosomatic symptoms | 0.08 [-0.17, 0.33] | 0.10 [-0.22, 0.41] | 0.18 [-0.03, 0.39] |

*Note.* Entries are standardised coefficients with 95% confidence intervals for departures from the 2023 Grindavík-national gap after the national time trend. Models adjust for expected grade cohort, gender and home language, with school-clustered CR2 standard errors; prior movers are excluded in 2025 and 2026. Each outcome is standardised within its restricted model sample, so these coefficients do not share the main benchmark's metric. The 2023 Grindavík series is one school and includes students later classified as prior movers; the departure intervals are conditional on that starting point. Home-language missingness and the grade-structured availability of belonging restrict the 2026 samples. In life-satisfaction/belonging/symptom order, analytic n was 18,423/18,573/18,469 in the three-wave models and 20,960/21,086/21,017 in the two-wave models.

The symptom items cover headache, stomachache, backache, dizziness, feeling low, irritability or bad temper, nervousness, and sleeping difficulties. Where six response categories were used, rarely and never were combined into five categories. Absence responses range from 1 (never) to 4 (more than three times). Valid HBSC substance-use responses are integers 1–7: 1 indicates never and 2–7 indicate use. Otherwise, valid IYS responses are used: 1 indicates yes and 2 indicates no. Any observed use counts as use; students with no observed substance items are missing.

Composition checks averaged evacuee–national contrasts from the boy/girl models over the fixed 2025 evacuee gender–grade mix, each wave's observed evacuee mix, and the fixed 2025 national-reference mix. The national-reference origin distribution conditional on gender and grade was held constant. Intervals condition on the target weights; absenteeism contrasts are risk differences. The life-satisfaction change was B = 0.17, 95% CI [-0.03, 0.36], under the fixed evacuee target and B = 0.21, 95% CI [0.01, 0.40], under the national target. These checks do not correct selective nonresponse.

**Table S8**
*Departures from the preregistration*

| Registered | In this paper |
|---|---|
| One 2025–2026 model with evacuees, prior movers, and the national reference group. | Evacuees are compared with the national reference group first. Prior movers are a separate model. |
| Gender as a three-group model by wave. | Gender is estimated for evacuees versus the national group. Prior-mover gender contrasts are a supporting check. |
| The 2023 benchmark emphasised school belonging and symptoms. | Life satisfaction is included as well. |
| Absenteeism, bullying, and substance use were not registered outcomes. | Absenteeism and substance use added for 2025–2026. Substance use promoted to a primary outcome after inspecting initial comparisons. Bullying reported in the supplementary material regardless of significance. |
| School-clustered HC2 standard errors. | Absenteeism and substance-use models use school-clustered HC2 standard errors. |
